# Mitigating isolation: The use of rapid antigen testing to reduce the impact of self-isolation periods

**DOI:** 10.1101/2021.12.23.21268326

**Authors:** Declan Bays, Timothy Whiteley, Matt Pindar, Johnathon Taylor, Brodie Walker, Hannah Williams, Thomas J. R. Finnie, Nick Gent

## Abstract

Isolating, either enforced or self-guided, is a well-recognised and used technique in the limitation and reduction of disease spread. This usually balances the societal harm of disease transmission against the individual harm of being isolated and is typically limited to a very small number of individuals. With the widespread transmission of SARS-CoV-2 and requirements to self-isolate when symptomatic or having tested positive, the number of people affected has grown very large causing noticeable individual cost, and disruption to the provision of essential services. With widespread access to reliable rapid antigen tests (also known as LFD or LFTs), in this paper we examine strategies to utilise this testing technology to limit the individual harm whist maintaining the protective effect of isolation. We extend this work to examine how isolation may be improved and mitigate the release of infective individuals into the population caused by fixed time-periods.

## Introduction

Interrupting the transmission of COVID-19 has been key in limiting disease spread in the community and reducing pressure on health care services. The use of non-pharmaceutical interventions (NPIs) has played a critical role in the public health responses adopted by governments throughout the COVID-19 pandemic. We have seen that policies which impose a period of self-isolation on confirmed cases and recent contacts of confirmed cases have proved effective at reducing onward transmission, and therefore also lessening the number of more acute infections. These policies are however subject to complications and limitations, and naturally incur an economic cost (1). As such, when considering social interventions, it is important that one balances the short-term cost in productivity against the longer-term gains acquired through reduced transmission. We strive to avoid total economic shut down, but need to appreciate the detrimental effect on mental and physical well-being caused by extended periods of self-isolation (2, 3).

Guidance on self-isolation periods provided by the European Centre for Disease Prevention and Control (ECDC) and World Health Organisation (WHO) initially suggested a 14-day isolation period for any close contact of a confirmed case (4, 5). This was reduced to 10 days in the summer of 2020 (6, 7); some European nations opted to reduce this again, adopting a 7-day isolation period in September 2020. This was motivated by a deepening understand of the epidemiological characteristics of SARS-CoV-2, specifically by evidence that transmission is rare after day 7 of infection (8). The current UK guidance requires confirmed cases to self-isolate for 10 days from the receipt of a positive test result. Additionally, fully vaccinated individuals who have come into recent contact with a confirmed case are strongly advised to undertake rapid antigen testing every day for 7 days thereafter, isolating if any of these return a positive result (correct as of 21/12/2021) (9).

The recent emergence of the Omicron variant (10) has seen case numbers increase dramatically, both internationally and domestically. With this, we see an increase in the risk of debilitating health and infrastructure services through excessive periods of self-isolation. Considering this, we present in this work an alternative strategy to manage individuals who test positive for SARS-CoV-2; utilising rapid antigen tests to facilitate an earlier end to the default isolation period. We do not consider PCR testing in this work as the turn-round time between swabbing and result makes it impractical for the purposes envisaged.

## Background

### Impact of isolation (positive cases) on the workforce

Across the working population, the numbers of available staff have been subject to fluctuation because of imposed self-isolation, causing an increased workload to be placed upon non-isolating staff or disruption to service provision. In the healthcare sector, academic staff have been required to work in the NHS full-time, and research fellows have returned to wards and clinics to meet demands (11), inevitably creating a backlog of work that the health service is not equipped to handle (12).

A survey conducted by the Royal College of Physicians in April 2020, at the peak of the first wave, determined that approximately 20% of healthcare staff were currently isolating due either to themselves or a household member developing COVID-19 symptoms (11). Another study of healthcare workers, determined that 44% of staff were required to self-isolate during April-July 2020 in response to developing COVID-19 symptoms, and 18% due to a household member developing symptoms (13). Additionally, a UK based survey of healthcare workers revealed that 28.9% of those surveyed had to self-isolate at least once due to developing COVID-19 symptoms or providing a positive test. Due to this, it was estimated that 11,800 – 21,000 working days were lost between February-May 2020. This equates to 71-127 working days lost per 1,000 health care workers (assuming a 40-hour working week) (14).

This reduction in staff has resulted in teams needing to revaluate risk-benefit scenarios to optimise pass-through rate, as opposed to providing care based on individual needs (11-13). In Oncology departments for example, regimens with less intensive treatments were being favoured in order to reduce the average bed-occupation time. Similarly, in radiotherapy, using fewer fractions with a higher dose per fraction was being considered, reducing the duration of treatment while simultaneously increasing the risk of toxicity (11).

Furthermore, healthcare workers from nonemergency, surgical allied health, community, and academic posts were having to be redeployed in order to meet the increase in demand that resulted from these pressures. Such actions also have consequences which will continue to be felt into the future, with disruption of services such as cardiology meaning fewer surgeries being performed and with it, the training capacity of new cardiac surgeons being greatly decreased.

In universities, students have found it difficult to adjust to isolation, leading to more students dropping out than usual. Staff have reported that students would refuse to participate in asymptomatic testing for fear of repercussions, including being made to isolate in the event of a positive test. University staff report “firefighting” through the pandemic, i.e., attempting to ease the burden of isolation on the student population, while simultaneously adhering to government guidelines (12).

The burdens faced by key workers not based in healthcare roles mirrors the burdens faced by the healthcare system. Self-isolation often resulted in staff shortages, increasing workloads and shift times for staff able to go into work – often without an increase in pay as compensation (15). Some companies have refused to take on adjusted timelines for projects, causing an increased pressure on staff (16). Additionally, staff have been required to take on tasks that they are unprepared and untrained to take on, resulting in potentially unsafe working conditions (16), additional stress/reduced wellbeing (15, 16) and increased workplace tensions amongst staff.

### Models of isolation regimes

Studies have quantified the effect duration of quarantine/ self-isolation has on COVID-19 spread. Ashcroft et al. used mathematical modelling to explore the effect of isolation duration in both confirmed SARS-CoV-2 cases as well as returning travellers. The role of strict isolation, as well as isolation with test and release strategies was investigated. Their modelling suggests the optimal ratio between fraction of transmission prevented and the number of days spent in quarantine with respect to societal and economic cost is delivered with an isolation duration of 7 days. For every 1 day increase in isolation, a reduction in onwards transmission is seen. However, these gains are considered marginal past day 10. The authors also concluded that the use of rapid antigen testing at day 5 with release on day 7 had similar efficacy as a test on day 6 followed by immediate release from isolation (82.3% CI: 68.2%, 93.4%) and (80.5% CI: 67.9%, 88.7%) respectively, meaning a shorter isolation period of 6 days with the use of rapid antigen testing before release is feasible (17).

Peng et al. investigated the effectiveness of a reduced COVID-19 quarantine time-period using a publicly available outbreak simulator (18). The authors drew several conclusions from the model; using RT-PCR testing 1-2 days before the end of a 10-day self-isolation period outperformed a 14-day test free self-isolation period. To achieve a post quarantine transmission risk (PQTR) of 0.1%, comparable to a 14-day self-isolation period, a 10-day period with one QT-PCR test or 2 rapid antigen tests can be used. Similarly, a reduced duration of 6 days can be achieved with the use of QT-PCR on days 4,5 and 6 whilst even shorter durations can be achieved using the higher PQTR of 1%, which is comparable to a 10-day, test free isolation period (19).

Quilty et al. used an agent-based model to simulate viral load dynamics of exposed contacts and their onward transmission potential in different quarantine and testing strategies. Assuming moderate levels of adherence to quarantine and self-isolation on symptom onset, self-isolation alone can prevent 35% of onwards transmission. Post exposure quarantine of 14 days reduces onwards transmission by 48% (95% UI 18-79). Self-isolation with release after a negative PCR test 7 days after exposure yields comparable results (50%, 95% UI 23–80; risk ratio [RR] 1·02, 95% UI 0·88– 1·41) to that of the 14-day self-isolation period. Isolation with a negative rapid antigen test 7 days after exposure (49%, 95% UI 20–78; RR 1·00, 0·82–1·28) or daily rapid antigen testing without quarantine for 5 days after tracing (50%, 95% UI 24–79; RR 1·04, 0·69–1·79) also yields similar efficacy (20).

The current literature indicates the feasibility of a shorter isolation period, especially in conjunction with multiple negative test results as a condition of release. These studies concur that a 7-day period with multiple tests yields similar onwards transmission to a 14-day isolation period with no testing and a 10-day isolation with testing, whilst reducing the economic and societal burdens of a longer isolation period. However, in a pilot study, close contacts of confirmed COVID-19 cases were given the option to carry out daily rapid antigen test as an alternative to self-isolation in the UK in December 2020. The participants were surveyed at the end of the study. Particularly noteworthy is that 13% of those who took part reported that they increased contacts following a negative test result (21). Although not directly related to positive cases, this study does highlight the risk of increased interactions following a single negative test result.

## Method

### Model and parameterisation

The model used within this paper is loosely based on the methodology presented in Bays et al (22). We use a Monte Carlo based model to simulate the process of 500,000 infected individuals being infected, identified and admitted into self-isolation in a given period. Depending on the scenario being considered, the isolated individuals may then undergo regular rapid antigen testing. Under some scenarios considered, a single negative test (whether true or false negative) will be sufficient to release individuals from self-isolation early. In others, we may require two consecutive negative tests. Individuals are assumed to be released at the end of their self-isolation regardless of infectious status.

Testing is assumed to produce a negative result for any individual who is tested post the end of their infectious period. For those who are still within their infectious period, test results are evaluated using a “weighted coin-toss”, where this weight is sampled according to the distribution provided in Table 1 for each individual. Each evaluation of our model will generate 100 artificial populations, each consisting of 500,000 infected individuals which are assumed to have been placed into isolation. We consider each of these populations separately to obtain confidence intervals on the values reported. For each simulated individual, we use the parameterisations given in Table 1 to sample an isolation start time, disease recovery time and rapid antigen test sensitivity. We have deliberately neglected test specificity due to current evidence suggesting this is very close to 100% (23), and omission will provide worst-case estimates for re-admission of infected individuals into the population. We assume the window of true rapid antigen test positivity coincides exactly with infectiousness (23).

**Table 1:** Parameter values and references used in the model

| Parameter | Distribution drawn from | Reference |
| --- | --- | --- |
| **LFT sensitivity** | Uniform (lower=0.7, upper=0.8) | (24) |
| **Infectious period distribution** | Gamma (shape=IPD_shape, scale=IPD_scale) | (25) |
| **IPD_shape** | Normal (mean=2.0, s.d.=0.1) | (25) |
| **IPD_scale** | Normal (mean=2.1, s.d.=0.1) | (25) |
| **Isolation entry** | Normal (mean=0, s.d.=0.3) | N/A |

Lastly, we assume that time t = 0 corresponds to the moment that each individual has fully incubated (i.e., when they would first start to test positive when tested). We define a ‘day’ as a 24-hour period. As individuals progress through our simulation, the model tracks the proportion which would be incorrectly released, how many hours on average each incorrectly released individual will remain infectious following release, and the average number of excess hours spent in isolation according to each of the self-isolation scenarios described below.

### Scenarios

Within this work we are interested in exploring a range of scenarios which might work to reduce excess isolation in the population. As such, we do not consider all permutations of isolation and testing. Instead, we look at only those that reasonably stand a chance of easing these pressures and being adopted as a policy. We describe these scenarios below. Note that in all scenarios, individuals are released upon completion of their isolation period regardless of infection status:

- 7, 10, and 14-day isolation with no pre-release testing.
- 10-day isolation with testing administered on days 7-9. Early release if a single negative result is returned.
- 10-day isolation with testing administered on days 6-9. Early release if two consecutive negative results are returned.
- 14-day isolation with testing administered on days 6-13. Early release if two consecutive negative results are returned.

To aid understanding, possibile pathways that an individual can proceed along under the third scenario of requring a double-negative test to end their isolation period eariler are demonstrated in Table 2. If they reach the end of the isolation period due to positive tests, then they are released regardless of whether they have had negative tests or not.

**Table 2:** Potential release scenarios under 10-day isolation and double-negative test scenario

| Day 6 | Day 7 | Day 8 | Day 9 | Day 10 |
| --- | --- | --- | --- | --- |
| -ve | -ve (release) |  |  |  |
| +ve | -ve | -ve (release) |  |  |
| -ve | +ve | -ve (do not release) | -ve (release) |  |
| +ve | +ve | -ve | -ve(release) |  |
| No consecutive negative rapid antigen tests |  |  |  | Release as usual |

## Results

Taking mean values, the proportion of people who are still infectious on each day is given in Table 3. This recapitulates earlier work (22) but provides a touchstone against which the reader may observe the self-isolation scenarios.

**Table 3:**
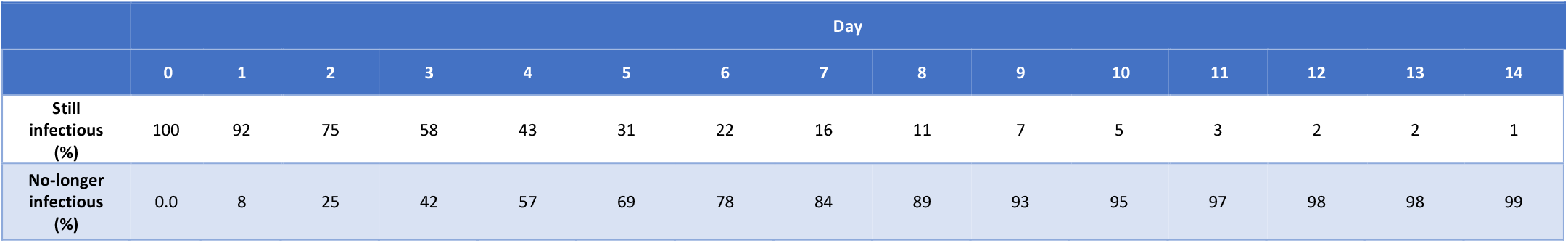
Percentages (rounded to integer) of people who are still infectious after each day according to their disease profile.

Table 4 then provides the major output from the model against the scenarios considered.

**Table 4:** Output from the model of the effect of the considered scenarios on disease release into the community (measured in three different ways) and the self-isolation in addition to that which is necessary to contain disease spread. Intervals are the 2.5 and 97.5 quantiles from the simulations.

| Policy | Released infectious (%) | Mean time a released person is infectious for (hours) | Mean excess isolation per person (hours) |
| --- | --- | --- | --- |
| 7-day isolation | 15.8% [11.9 - 21.0] | 62.3 [56.5 - 69.2] | 76.8 [67.2 - 84.0] |
| 10-day isolation | 5.1% [3.4 - 7.6] | 59.3 [53.5 - 65.6] | 141.6 [129.6 - 151.2] |
| 14-day isolation | 1.0% [0.6 - 1.8] | 57.1 [51.4 - 63.1] | 235.2 [220.8 - 247.2] |
| **10-day isolation, or 1 negative tests from day 7** | 9.2% [6.5 - 12.8] | 61.1 [55.3 - 67.5] | 79.2 [69.6 - 86.4] |
| **10-day isolation, or 2 negative tests from day 6** | 6.2% [4.2 - 9.0] | 60.0 [53.9 - 66.3] | 81.6 [72.0 - 88.8] |
| **14-day isolation, or 2 negative tests from day 6** | 4.1% [2.6 - 6.0] | 61.3 [55.8 - 67.7] | 69.6 [64.8 - 74.4] |

## Discussion

### Overview

The current 10-day isolation period results in the release of 5% of the infected population being released from isolation whilst still being infectious. This reduces to 1% when a 14-day isolation period is considered. In comparison, 10-day isolation including 2 rapid antigen test negative results from day 6, provides a large reduction in excess isolation in return for a minimal cost of releasing those who are still infectious. Under this regime, 6% of people are released infectious. Excess isolation is reduced from 142 hours to 82 hours.

The optimal solution mathematically is unlimited mandatory isolation, which people can leave once they are no longer infectious. This is not practically possible as people’s true disease status is unknowable, false negative and positive rates from tests, and the likelihood that some individuals with unusual biology (for example the immune-supressed) may never show negative. Under the considered options we introduce a ceiling on isolation time to the originally recognised 14 days and assess the course of the disease from day 7 onward with consecutive lateral flow tests. This reduces both the average time spent isolating unnecessarily and the time that people are released whilst infectious.

Care must still be exercised for the period following someone’s release from isolation as in all scenarios there is a risk of releasing an infective person. A 7-day isolation period alone is a notably poor solution as 16% of people could be released prematurely. A single negative rapid antigen test also does not appear sufficient to end isolation because there is still risk of a false negative and a 9% chance of premature release.

### Exploring 10 days isolation

We now look in more detail at when people would be released from isolation and the proportions of those being released correctly and falsely at each step. The breakdown according to the day of release can be seen in Table 5.

**Table 5:** Breakdown of the “10-day isolation, or 2 negative rapid antigen tests from day 6” policy. The majority of people (79%) are released correctly on day 7. Conversely, there are a significant minority who end their isolation yet are still infectious (4.2%). Intervals are the 2.5 and 97.5 quantiles from the simulations.

|  | Day 6 | Day 7 | Day 8 | Day 9 | Day 10 | End of isolation |
| --- | --- | --- | --- | --- | --- | --- |
| **New false releases** | 0 [0 - 0] | 1.0 [0.6 - 1.5] | 0.5 [0.3 - 0.7] | 0.3 [0.2 - 0.5] | 0.2 [0.1 - 0.3] | 4.2 [2.5 - 6.4] |
| **New true release** | 0 [0 - 0] | 79.0 [73.2 - 84.2] | 6.0 [5.0 - 7.0] | 4.3 [3.4 - 5.3] | 2.9 [2.1 - 3.7] | 1.6 [1.1 - 2.2] |
| **Previously released** | 0 [0 - 0] | 0.0 [0.0 - 0.0] | 80.0 [74.1 - 84.9] | 86.5 [81.6 - 90.4] | 91.2 [87.3 - 94.0] | N/A |
| **Still in isolation** | 100 [100 - 100] | 20.0 [15.1 - 25.9] | 13.5 [9.6 - 18.4] | 8.8 [6.0 - 12.7] | 5.8 [3.7 - 8.6] | N/A |

We can see that most people (79%) will be released on day 7 of their isolation. Only 6% of people will make it to the full 10 days of isolation. However, the majority of those who made it to day 10 would still need to isolate for even longer.

The biggest source of ‘false releases’ is caused by releasing people after 10 days. 2% of people will be falsely released based on testing as opposed to 4% because of the end of the 10-day isolation period.

### Comparing mandatory isolation days

Lastly, we look at different days from which people begin testing. We plot the percentage of false releases in each case in Figure 1. This figure accounts for all the people who leave isolation at different points and does not necessarily mean people will leave after the minimum isolation period has ended but when they have had two negative rapid antigen tests.

**Figure 1:**
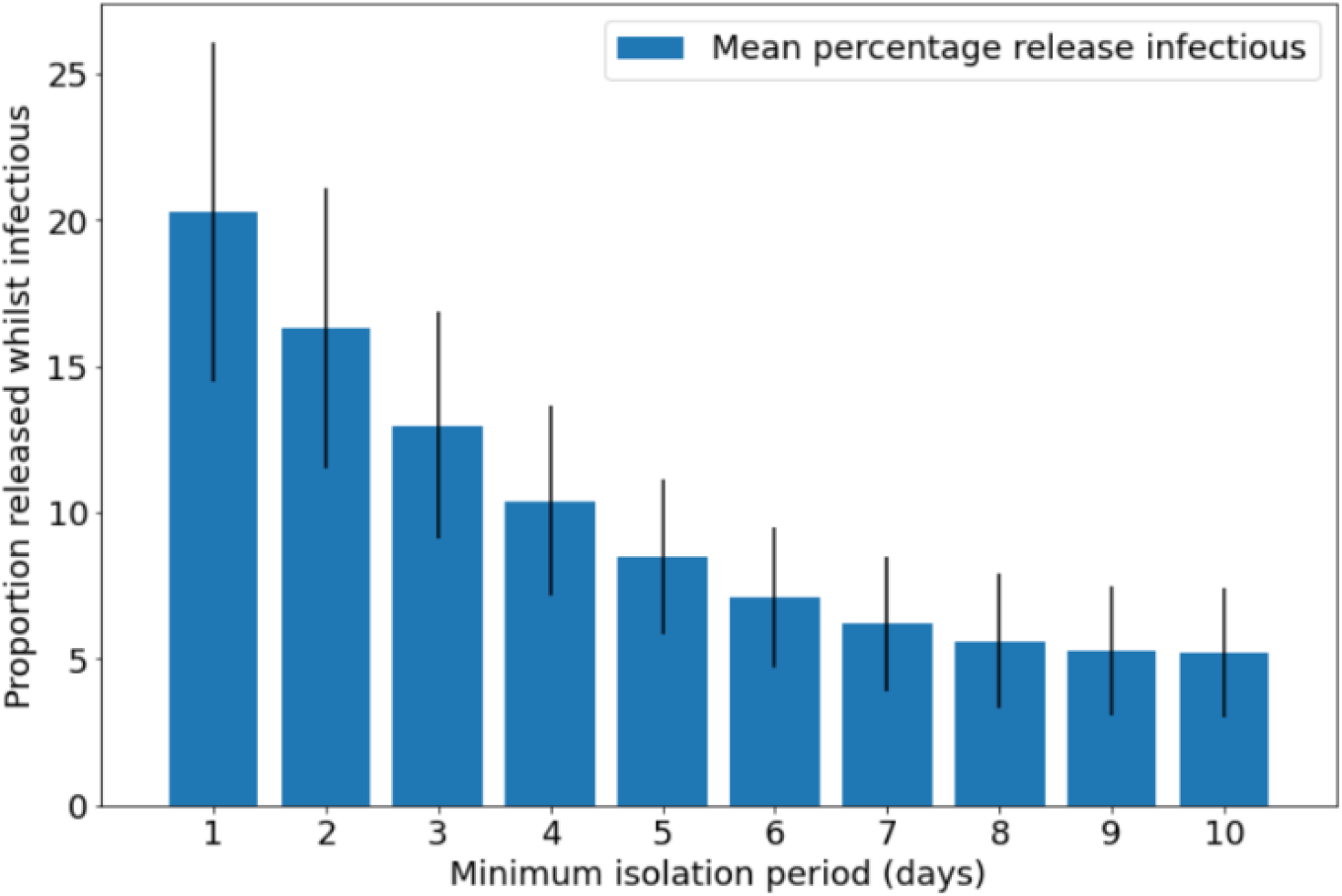
Sensitivity of the model to different testing regimes. People will not necessarily be released on the first day possible but when they have two consecutive negative LFD tests, or when they reach 10 days. We are measuring the cumulative effects over all releases over all the days

We can see that removing a mandatory isolation period and relying only on 2 rapid antigen tests alone appears unwise due to the risk of false negatives. The risk of which will increase with the number of tests taken, and hence increase over time. A mandatory isolation period of longer than 7 days will not provide much more safety but lowering this mandatory isolation point will increase the percent of ‘false releases’ dramatically.

#### Limitations

We have not considered compliance in this model. This is explicit as compliance is a complex and multifaceted behavioural science problem which is far beyond the scope of this simple physical-system model.

Secondly, while the sensitivity of rapid antigen tests is well known within laboratory conditions, quantifying this in real-world situations is not trivial. To overcome this, we have incorporated some uncertainty into the model by using a random variable based on real-world conditions to describe the rapid antigen test sensitivity assigned to each simulated individual.

Lastly, we have considered time within this system from a purely mathematical viewpoint. That is, days are 24-hour periods and the testing/releasing of simulated individuals occurs exactly at unit periods. In conversion to the real-world we understand that should someone begin what they consider day 0 at 23.59 and conducts their tests at 07:00 this will shorten the time window compared to the model. Figure 1 shows the results are still reasonably robust in this scenario however care is required in translation of time periods.

## Conclusion

With this modelling work we can see that there is a way to reduce the period of self-isolation required to prevent disease transmission with the use of high-specificity, rapid antigen testing.

Within the bounds of current UK guidance, taking rapid antigen tests from day 6, and requiring 2 consecutive negative tests 24 hours apart, a regime is generated that would release 79% of people correctly on day 7, with 6% of people requiring to stay in isolation until day 10. The total percentage of people released whilst still infectious will be approximately equivalent whereas the excess isolation time will drop from 6 days to 3 days. Note that it is key to this regimen that people should not end isolation early without the two negative rapid antigen tests as there is significant risk that they will still be infectious.

In the absence of available tests, the system should revert to a simple upper-bound on isolation period. Such a bound should be set dependent on risk appetite where, say, 5% of infected individuals would be released with 10 days of plain isolation and 1% with 14 days. In all cases, we urge caution as there is still a chance of residual infectiousness. In particular, should a person be still positive with a rapid antigen test at, for example, day 10 then we would encourage further isolation until two clear tests are obtained.

Outside of the current guidance the most beneficial scenario is one where we both reduce the mandated minimum isolation period but allow for an unconstrained maximum isolation period whilst evidence of infection is still present. Thus, by allowing the release of each individual based on their own disease course and hence risk profile. Practically, some constraint on isolation period may be desirable. In which case we have shown here that 14 days may provide a sensible upper bound.

## Data Availability

All data produced in the present study are available upon reasonable request to the authors

## Acknowledgements

The authors gratefully acknowledge the help and assistance of Emma Bennett in supplying the epidemiological evidence and for searches of the literature.

